# The proportion of deaths cases in confirmed patients of COVID-19 is still increasing for cumulative cases reported up to 25 April 2020

**DOI:** 10.1101/2020.04.17.20068908

**Authors:** Thomas Scheuerl

**Affiliations:** Research Institute for Limnology, Mondsee, University of Innsbruck, Mondsee, Austria

## Abstract

In this work I analyse how proportions of fatal cases after COVID-19 infection change since outbreak of the disease. Using publicity available data I model the change in deaths probability from day 30 of outbreak until 25 April 2020. The global trend is that the proportion of fatal cases is still increasing and that many countries have not yet reached the maximum deaths proportion. However, there are visual differences between countries and in some countries the proportions are clearly below or above the global trend. A positive correlation between deaths cases and recorded infections indicates that a higher infection number results in increased mortality numbers.

## Introduction

The ongoing coronavirus (COVID-19) outbreak starting in China [1] expanded rapidly over the globe and many countries respond to the spread of the disease by a lock-down to limit contacts between people. Many countries are heavily affected by the outbreak and there is a high deaths rate [2]. The deaths probability will be an important parameter guiding how long lock-down and social distancing are necessary. Countries economies are under pressure and for older adults, who are the most vulnerable group, social distancing is predicted to come with serious negative effects [3], but obviously spread of the disease should be condemned as much as possible until the health system can cope with the disease. Here, I analyse the death proportion dynamics of COVID-19 infections since outbreak of the disease. I focus on the change of deaths probability over confirmed cases of a particular day and if this proportion changes over time. This indicates the probability of a fatal case for the current number of cases and reflects whether further increase can be expected, or if proportions have reached the maximum. The analysis reveals that at the current number of confirmed cases, the proportion of people dying is still increasing in most countries, indicating the situation may further worsen.

## Methods

I analyse freely available data on the COVID-19 pandemic situation from Hopkins University https://coronavirus.jhu.edu/data/mortality. Data were downloaded from https://github.com/datasets/covid-19/tree/master/data. The data frame contains 185 countries and collects *confirmed cases, recovered cases* and *deaths cases* each day starting from 22^nd^ of January 2020. I limit the analysis to countries which have at least 1000 confirmed cases and countries that have at least 1 recovered and 1 fatal case. Because the disease was spreading in China several days before it arrived in other countries, I also removed the first 30 days, during which deaths cases were limited to China.

Data were analysed in R statistical environment [4]. A time-series analysis was conducted to track changes over time using generalized additive mixed effects models (GAMMs) following ref. [5]. This approach smoothes changes over time tracking non-linear trends. Change in proportion and count data were modelled using ‘Days’ as continuous variable building a global model for all countries applying a smooth term with family either *binomial* for proportion or *poisson* for count data. The change over time per country was introduced into the random effect. The models were build using the R packages *nlme* [6] and *mgcv* [7]. The change over time was smoothed using 10 knots and a *cubic regression* spline. Model convergence was controlled using the *lmeControl* function, with the maximum number of iterations set to 500 and using optimisation *opt=optim* function. Temporal autocorrelation was introduced to account for violation of independence. Choosing based on the lowest AIC criterion, the best correlation structure was selected from a range of possibilities presented elsewhere [5]. All final models included a *corAR1* correlation structure including time and country to address temporal correlation. Data were *log10* transformed to improve model fit, inspected by validation plotting. To explore the correlation of deaths proportions over confirmed cases generalized least square models were used including the *varExp* variance structure to allow changing spread of residuals over changes in confirmed cases. Visualization was also done in R.

## Results

There are >70 countries with more than 1000 confirmed cases and at least one 1 deaths and 1 recovered case.

The global trend for new cases is still increasing but slightly slowing down (Supporting Fig. 1). The smoothed term across time for all countries indicates an increase with little decrease. Only in China and South Korea no new cases are confirmed. In all other countries the trend is still raising, however not exponential any more. Most cases are confirmed in the US, followed by Italy, Spain and Germany, but in all countries the increase of new cases is slowing down. Japan is one of the countries following the global trend. Thailand is one of the countries with an increase less than the global trend in new cases.

There also is a positive trend for recovered cases. The smoothed term across all countries however indicates that this trend is starting to slow down. Not surprisingly, those countries with highest confirmed cases have most recovered cases.

The same seems true for deaths cases. In all countries, except China, the numbers of deaths cases are increasing daily (Supporting Fig. 3).

I also looked into a measure like “Open cases” by subtracting daily deaths cases and daily recovered cases from the total number confirmed (Supplemental Fig. 4). Using this kind of measure, global open cases are still increasing as indicated by the global smooth term. The countries China, South-Korea and Thailand seem to record a reduction of open cases, while in all other countries more open cases are recorded daily.

Up to this point there is little new information more than in most countries more and more cases are recorded. Next I was looking into how the temporal proportions change and if the likelihood to either die or recover is changing since outbreak of the disease. I start on looking at the proportion of deaths per confirmed cases. This measure may represent how likely deaths occurs and if the maximum is reached already. The global trend is still positive, indicating the proportion of deaths cases is still going to increase world wide based on total cases (Fig. 1). However, there is a decline indicated and there is no further significant increase the last few days.

**Figure 1.**
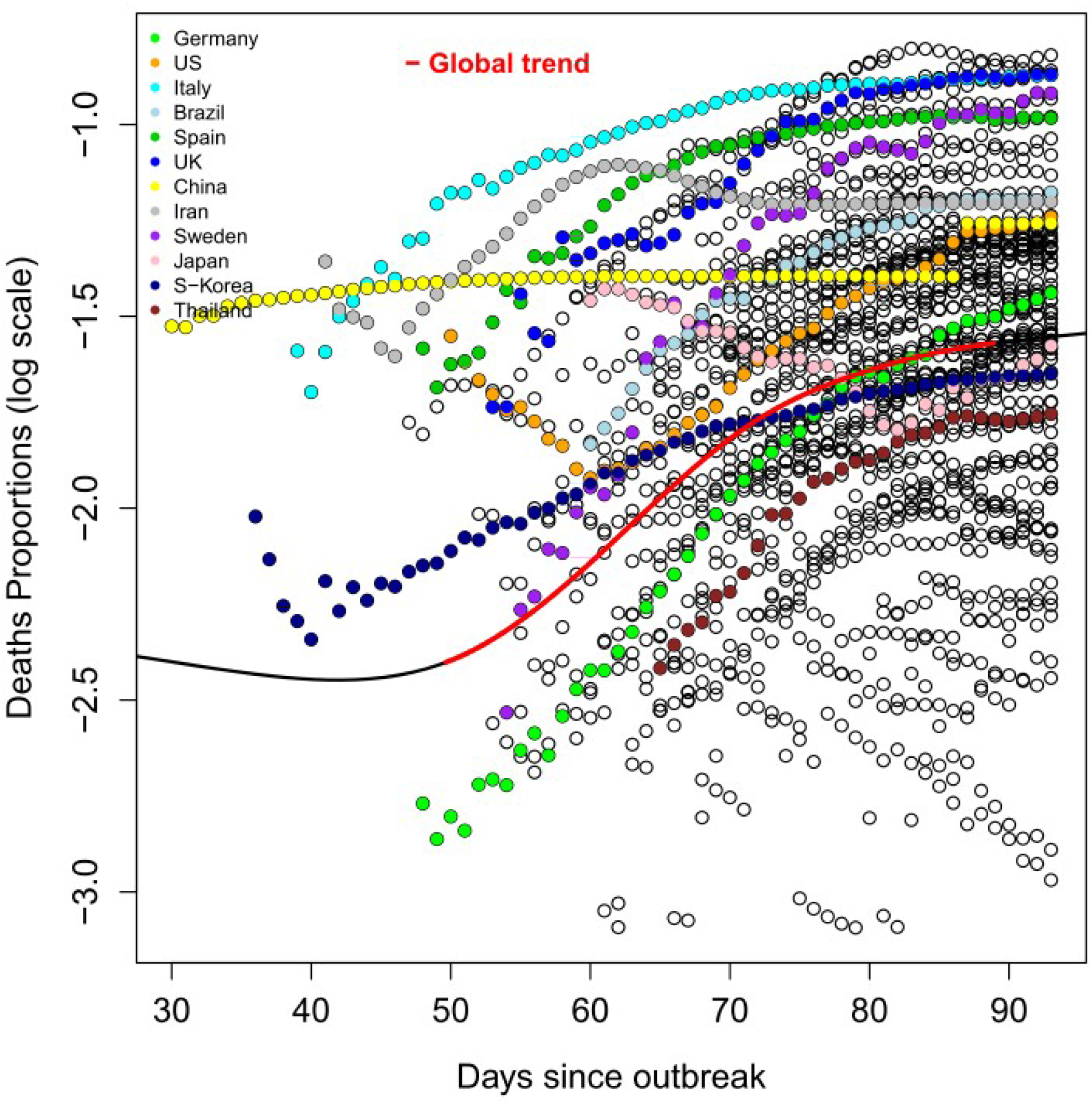
The proportion of fatal cases after COVID-19 infection since outbreak. The proportion is shown on a log scale. The global trend (black line) indicates an increasing trend (significant trend in red) with little reduction in daily new cases. Temporal tracks of individual countries are indicated by different colours as indicated in the legend.

In countries like Italy, Spain and the United Kingdom, the deaths proportions are particularly high, but now seem to level off and stabilise. In China this measure doesn’t change because there are no new cases of infections, neither deaths cases recorded (Supporting Figures 1+3), which causes no more temporal change. This indicates the maximum proportion was reached. Interestingly, in the country with most confirmed cases, the United States, the deaths proportions are lower than could be expected. Germany is reaching the global trend now, despite it has more than average confirmed cases. In Japan the deaths proportions are declining, which may be caused by the fact that there are more daily new infections than on average, but the deaths rates are increasing less than the global trend.1

One likely explanation for the observed patterns of increased deaths proportions over time after the pandemic outbreak may be that more cases simply result in even more deaths cases because the countries health system experiences an overload. Because of this I looked into correlations between confirmed cases and deaths proportion, checking for positive trends. Indeed there seems to be a positive trend that more confirmed cases result in a higher probability of fatal recordings (Fig. 2).

**Fig 2.**
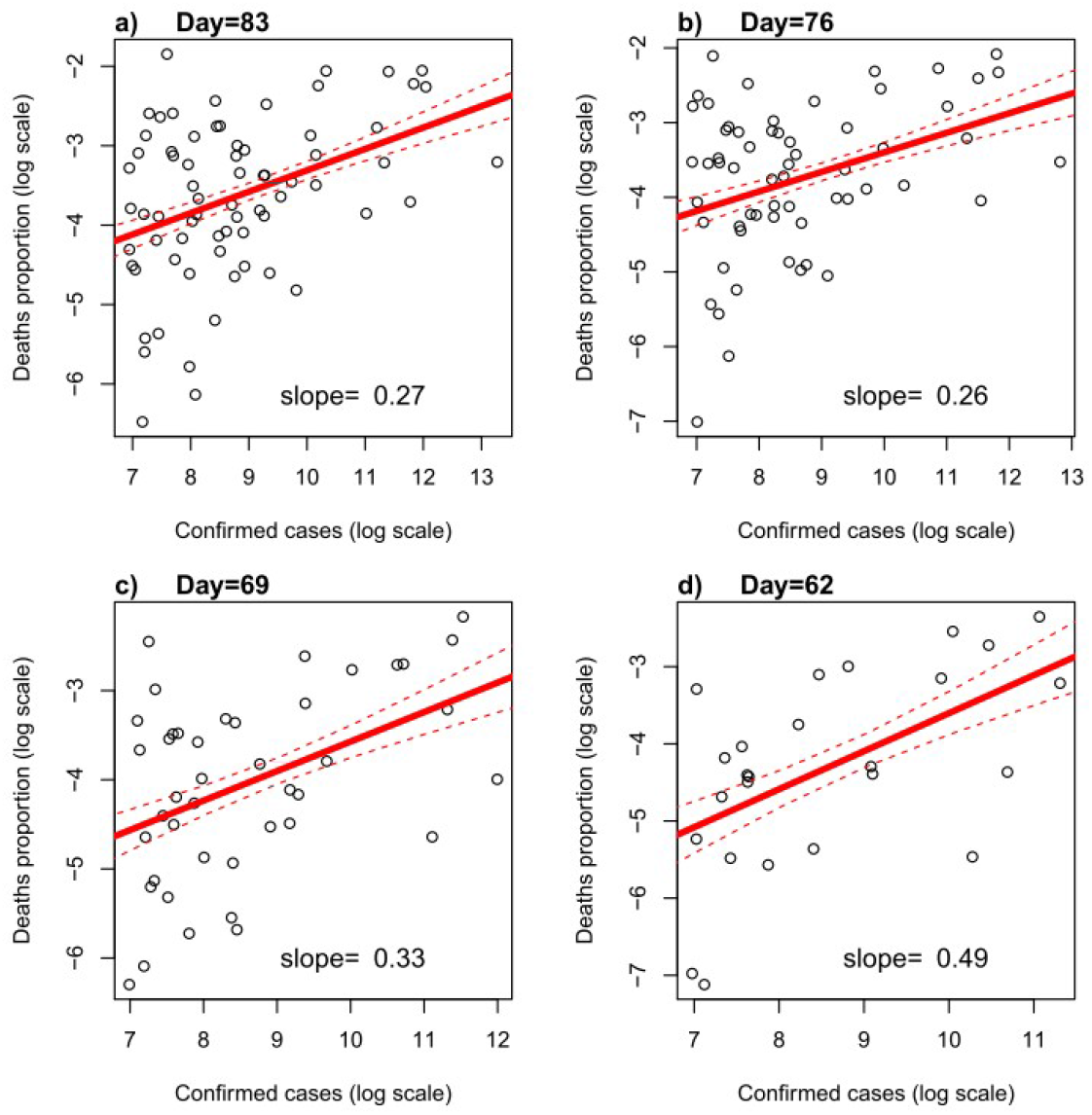
The correlation between deaths proportions and number of confirmed cases for four different days within the last four weeks. The red line indicates the best fit line of a generalized least square model including a variance structure allowing for larger residual spread for low number of confirmed cases. Individual slopes are indicated per plot.

With more infected cases the probability to die from COVID-19 infections seems to increase. This is true for 4 different days within the last month. In line with the global analysis the correlations are all positive but slowly decrease over time.

The proportions of recovered cases is also increasing however this trend seems also to be slowing down (Supporting Figure 5). Countries like the US, UK and Sweden seem to be below the global trend.

## Discussion

Coronavirus causes severe respiratory symptoms resulting in many fatal cases. The number of infections is still increasing in many countries and so do the proportions of fatal cases. Giving hope, there is a declining global trend that the proportions start to level off. However, there are clear differences between countries and individual countries will have to follow individual actions to condemn the spread of the disease. While in general it looks like more infections cause more fatal cases, this is not true or all countries. Particularly in the US the deaths proportions are less increasing than might be expected. This may be either that the healths system can still cope with the cases, or confirmed cases are accumulating faster than fatal cases rise.

The deaths over confirmed analysis is a bit inaccurate as when there are no records neither in new confirmed cases nor new deaths cases, like in China, there is no decline in the proportions. It only tells if the maximum is yet reached. In case of China the measure indicates the country has reached the maximum level in deaths probability. Another measure could be to calculate “Deaths cases” over “Open cases”. This would require to calculate daily new cases of deaths only. This is a promising future step and certainly worth following up. In this work I focus on whether countries have already reached the maximum deaths proportion and if this trend seems stabilizing over time.

## Data Availability Statement

All relevant data are within the paper and its Supporting Information files.

## Data Availability

All relevant data are within the paper and its Supporting Information files.

## Supplementary Figures

**Supplementary Figure 1.**
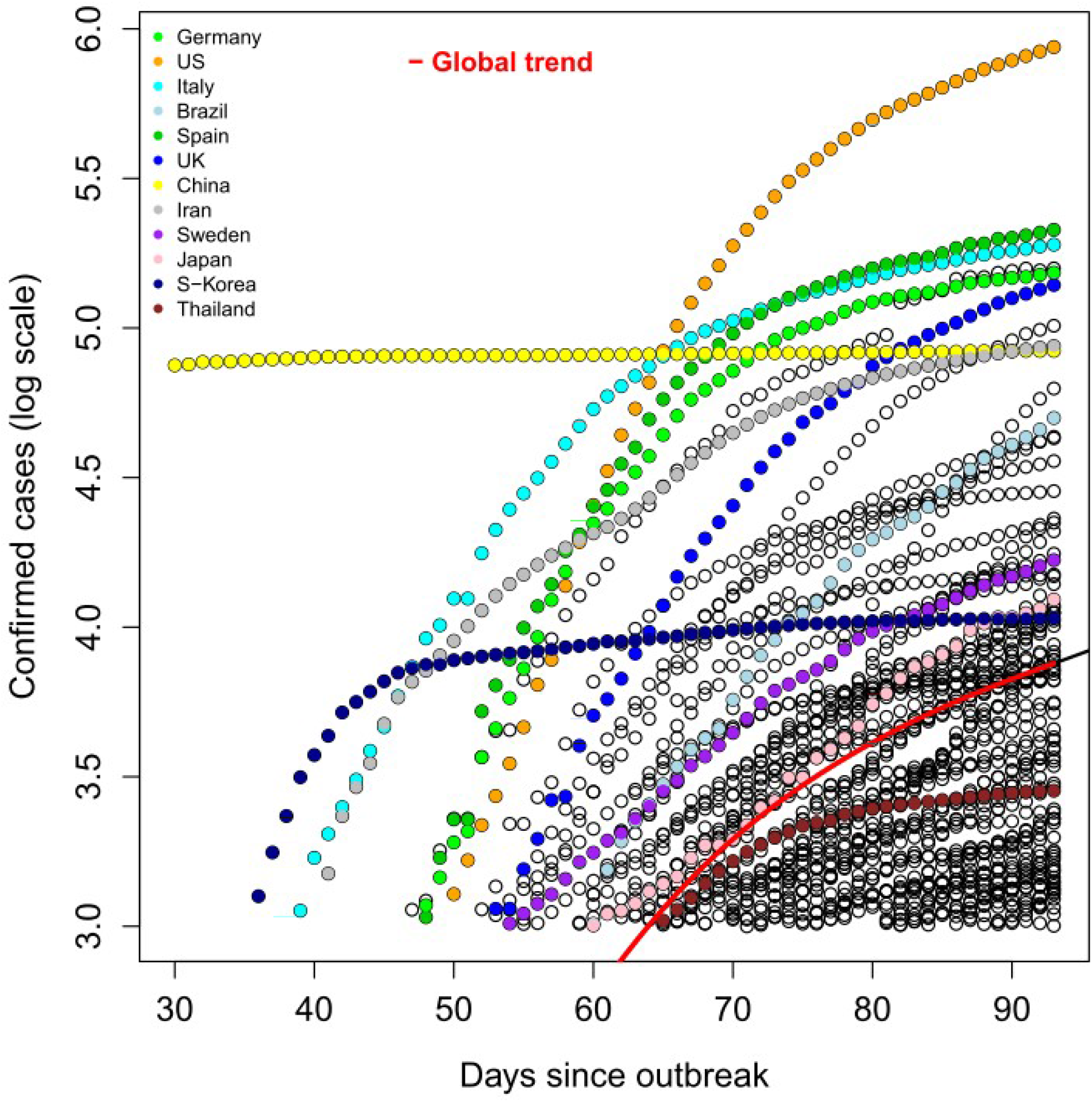
Confirmed new cases of COVID-19 since outbreak. The confirmed number is shown on a log scale. The global trend (red line) indicates an increasing trend with little reduction in daily new cases. Temporal tracks of individual countries are indicated by different colours as indicated in the legend.

**Supplementary Figure 2.**
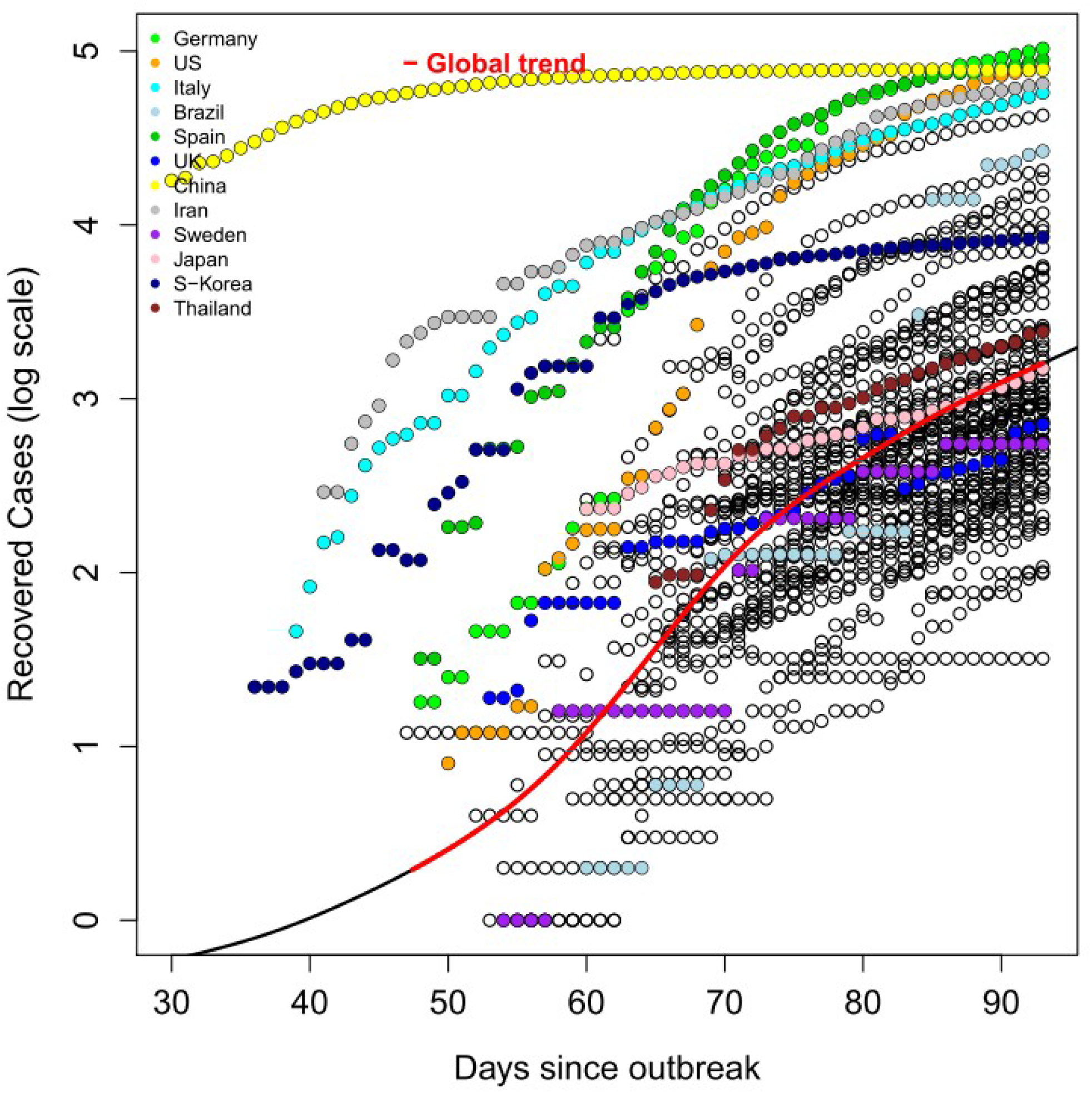
Recovered cases since outbreak. The recovered cases are shown on a log scale. The global trend (red line) indicates an increasing trend with little reduction in daily new cases. Temporal tracks of individual countries are indicated by different colours as indicated in the legend.

**Supporting Figure 3.**
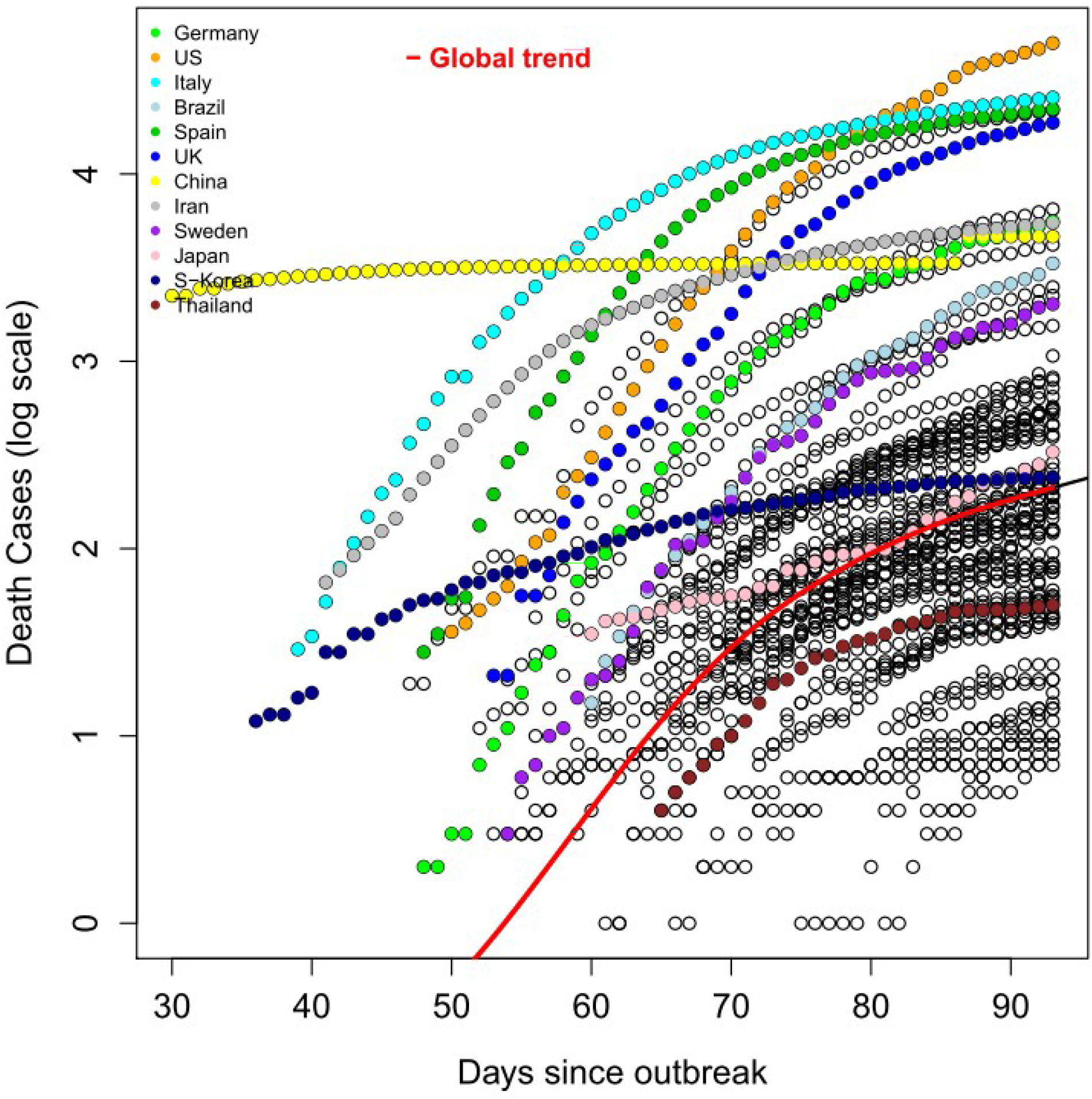
Deaths cases since outbreak. The dearths cases are shown on a log scale. The global trend (red line) indicates an increasing trend with little reduction in daily new cases. Temporal tracks of individual countries are indicated by different colours as indicated in the legend.

**Supporting Figure 4.**
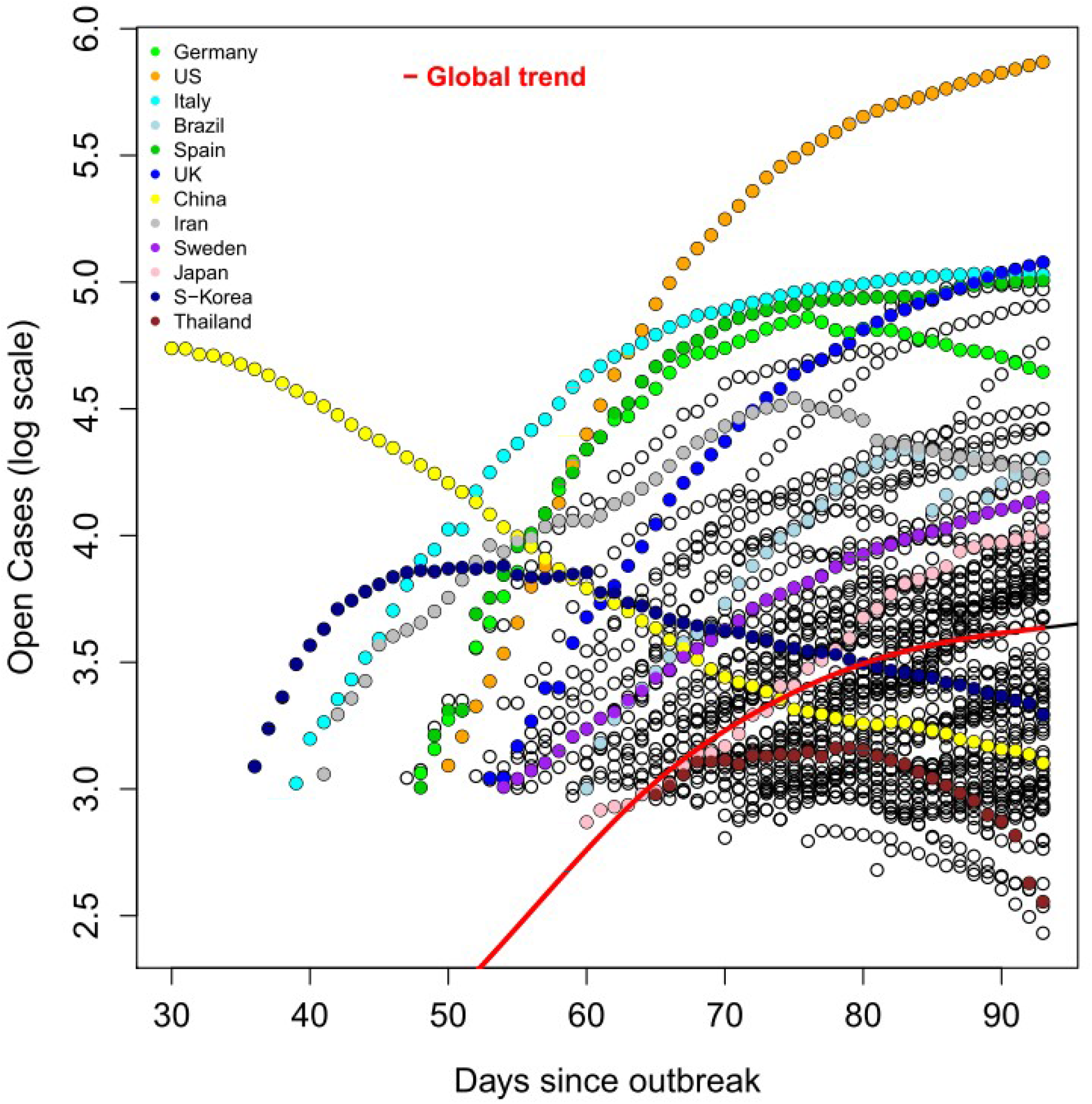
Open cases since outbreak. The open cases are shown on a log scale. The global trend (red line) indicates an increasing trend with little reduction in daily new cases. Temporal tracks of individual countries are indicated by different colours as indicated in the legend.

**Supporting Figure 5.**
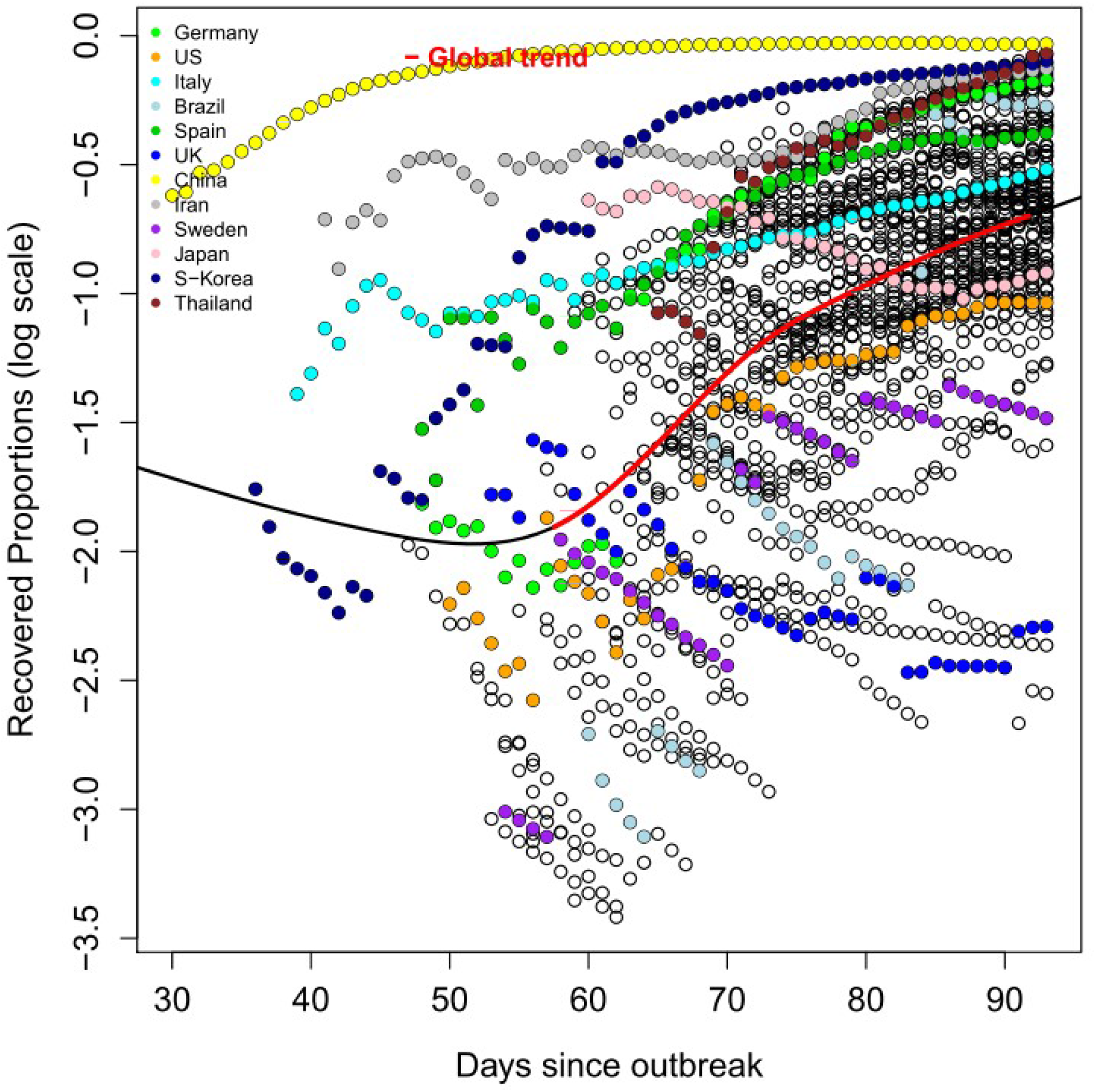
Proportion of recovered cases since outbreak. The proportions of recovered cases are shown on a log scale. The global trend (red line) indicates an increasing trend with little reduction in daily new cases. Temporal tracks of individual countries are indicated by different colours as indicated in the legend.

